# A high-resolution HLA reference panel capturing global population diversity enables multi-ethnic fine-mapping in HIV host response

**DOI:** 10.1101/2020.07.16.20155606

**Authors:** Yang Luo, Masahiro Kanai, Wanson Choi, Xinyi Li, Kenichi Yamamoto, Kotaro Ogawa, Maria Gutierrez-Arcelus, Peter K. Gregersen, Philip E. Stuart, James T. Elder, Jacques Fellay, Mary Carrington, David W. Haas, Xiuqing Guo, Nicholette D. Palmer, Yii-Der Ida Chen, Jerome. I. Rotter, Kent. D. Taylor, Stephen. S. Rich, Adolfo Correa, James G. Wilson, Sekar Kathiresan, Michael H. Cho, Andres Metspalu, Tonu Esko, Yukinori Okada, Buhm Han, NHLBI Trans-Omics for Precision Medicine (TOPMed) Consortium, Paul J. McLaren, Soumya Raychaudhuri

## Abstract

Defining causal variation by fine-mapping can be more effective in multi-ethnic genetic studies, particularly in regions such as the MHC with highly population-specific structure. To enable such studies, we constructed a large (N=21,546) high resolution HLA reference panel spanning five global populations based on whole-genome sequencing data. Expectedly, we observed unique long-range HLA haplotypes within each population group. Despite this, we demonstrated consistently accurate imputation at G-group resolution (94.2%, 93.7%, 97.8% and 93.7% in Admixed African (AA), East Asian (EAS), European (EUR) and Latino (LAT)). We jointly analyzed genome-wide association studies (GWAS) of HIV-1 viral load from EUR, AA and LAT populations. Our analysis pinpointed the MHC association to three amino acid positions (97, 67 and 156) marking three consecutive pockets (C, B and D) within the HLA-B peptide binding groove, explaining 12.9% of trait variance, and obviating effects of previously reported associations from population-specific HIV studies.

## Main

The HLA genes located within the MHC region encode proteins that play essential roles in immune responses including antigen presentation. They account for more heritability than all other variants together for many diseases^1–4^. It also has more reported GWAS trait associations than any other locus^5^. The extended MHC region spans 6Mb on chromosome 6p21.3 and contains more than 260 genes^6^. Due to population-specific positive selection it harbors unusually high sequence variation, longer haplotypes than most of the genome, and haplotypes that are specific to individual ancestral populations^7,8^. Consequently, the MHC is among the most challenging regions in the genome to analyze. Advances in HLA imputation have enabled population-specific association and fine-mapping studies of this locus^2,9–12^. But despite large effect sizes, fine-mapping in multiple populations simultaneously is challenging without a single large and high-resolution multi-ethnic reference panel. This has caused confusion in some instances. For example, defining the driving HLA alleles may inform the design of antigenic peptides for vaccines^13,14^ for HIV-1, which led to 770,000 deaths in 2018 alone^15^. However, multiple risk HLA risk alleles have been independently reported in different populations^1,10,16^, and it is not clear if they represent truly population-specific signals or are confounded by linkage.

## Results

### Performance evaluation of inferred classical HLA alleles

To build a large-scale multi-ethnic HLA imputation reference panel, we used high-coverage whole genome sequencing (WGS) datasets^17–21^ from the Japan Biological Informatics Consortium^20^, the BioBank Japan Project^18^, the Estonian Biobank^22^, the 1000 Genomes Project (1KG)^21^ and a subset of studies in the TOPMed program (**Supplementary Note, Supplementary Table 1-2**). To perform HLA typing using WGS data, we extracted reads mapped to the extended MHC region (chr6:25Mb-35Mb) and unmapped reads from 24,338 samples. We applied a population reference graph^23–25^, for the MHC region to infer classical alleles for three HLA class I genes (HLA*-A, -B* and -*C*) and five class II genes (HLA*-DQA1*, -*DQB1, -DRB1*, -*DPA1*, -*DPB1*) at G-group resolution, which determines the sequences of the exons encoding the peptide binding groove. We required samples to have >20x coverage across all HLA genes (**Supplementary Table 1, 3**). After quality control our panel included 21,546 individuals: 10,187 EUR, 7,849 AA, 2,069 EAS, 952 LAT and 489 SAS.

**Table 1.**
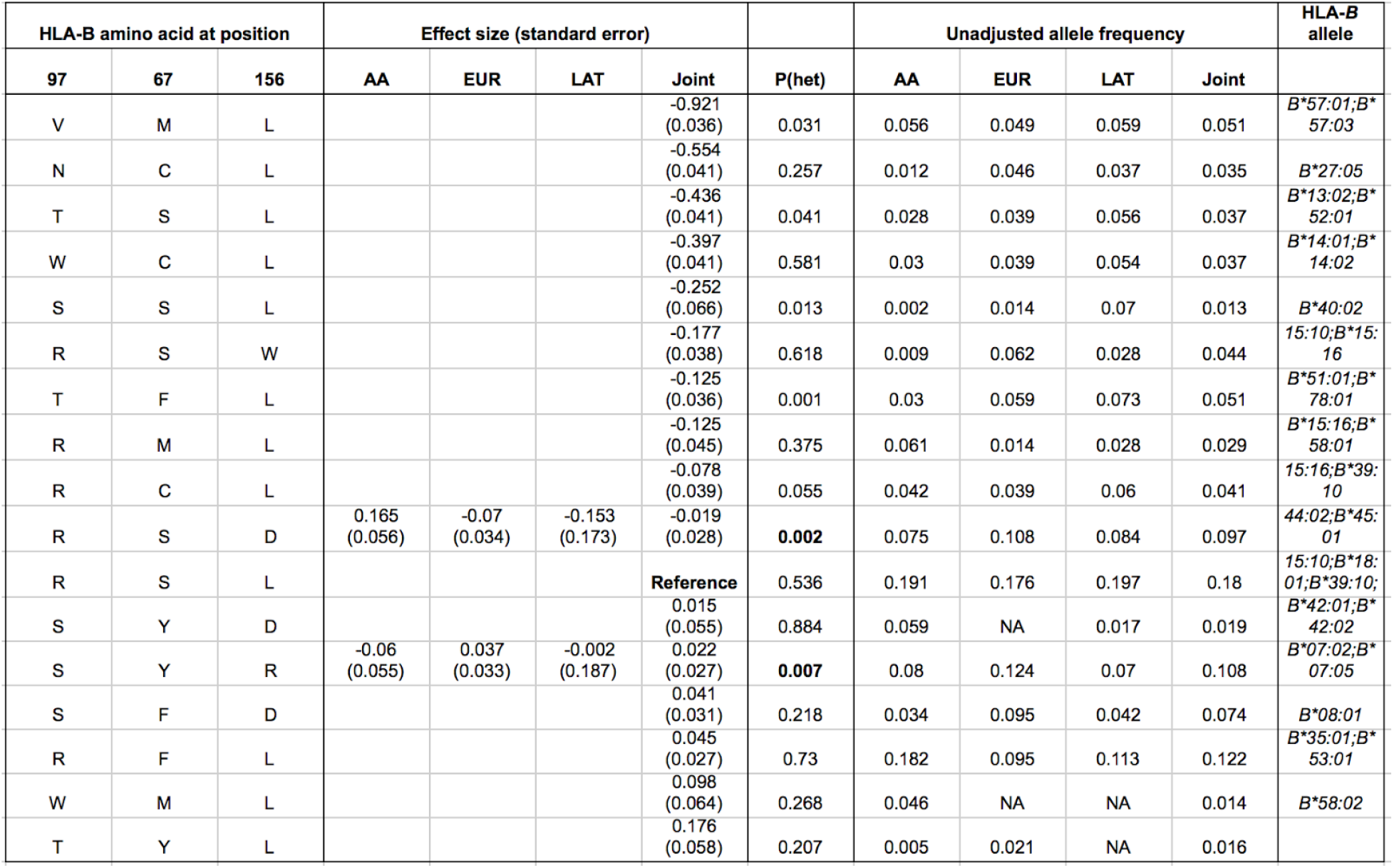
Effect estimates for the haplotypes defined by the three independent amino acids in HLA-B associated with HIV-1 viral load. Only haplotypes with >1% frequency in the overall population are listed (**Supplementary Table 15**). Classical alleles of HLA-B are grouped based on the amino acid residues presented at position 97, 67 and 156 in HLA-B. For each haplotype, the multivariate effect is given as an effect size, taking the most frequent haplotype (97R-67S-156L) as the reference (effect size = 0). Heterogeneity p-value (P(het)) of each haplotype is calculated using a F-statistics with two degrees of freedom (**Methods**). Effect size and its standard error in each population are listed only for haplotypes that show evidence of heterogeneity (P-value < 0.05 /26, bolded). Unadjusted haplotype frequencies are given in each population.

To assess the accuracy of the WGS *HLA* allele calls, we compared the inferred *HLA* classical alleles to gold standard sequence-based typing (SBT) in 955 1KG subjects and 288 Japanese subjects and quantified concordance. In both cohorts we observed slightly higher average accuracy for class I genes, obtaining 99.0% (one-field, formally known as two-digit), 99.2% (amino acid) and 96.5% (G-group resolution), than class II genes, obtaining 98.7% (one-field), 99.7% (amino acid) and 96.7% (G-group resolution, **Methods, Supplementary Figure 1, Supplementary Tables 4-5, Extended Data 1**).

**Figure 1.**
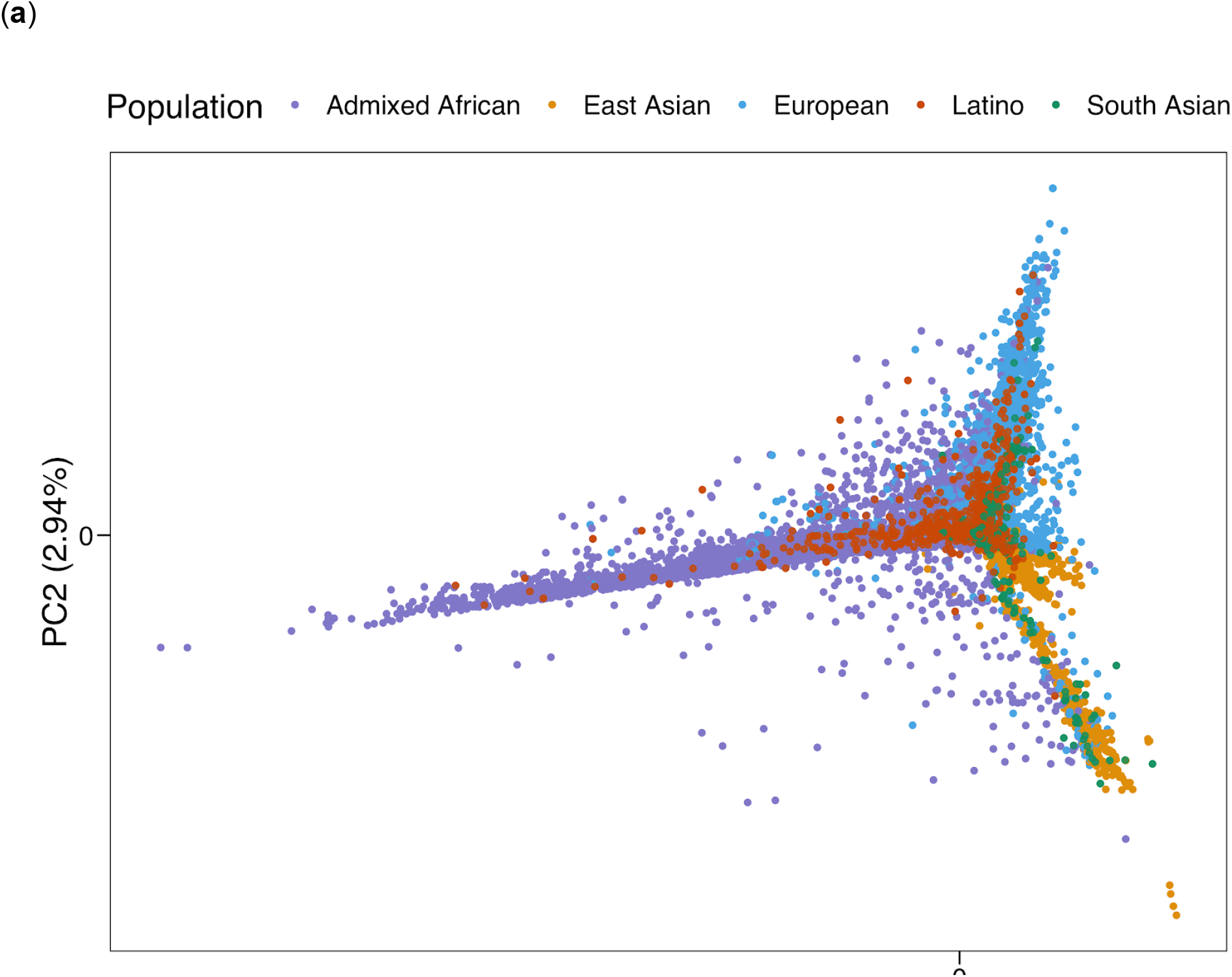
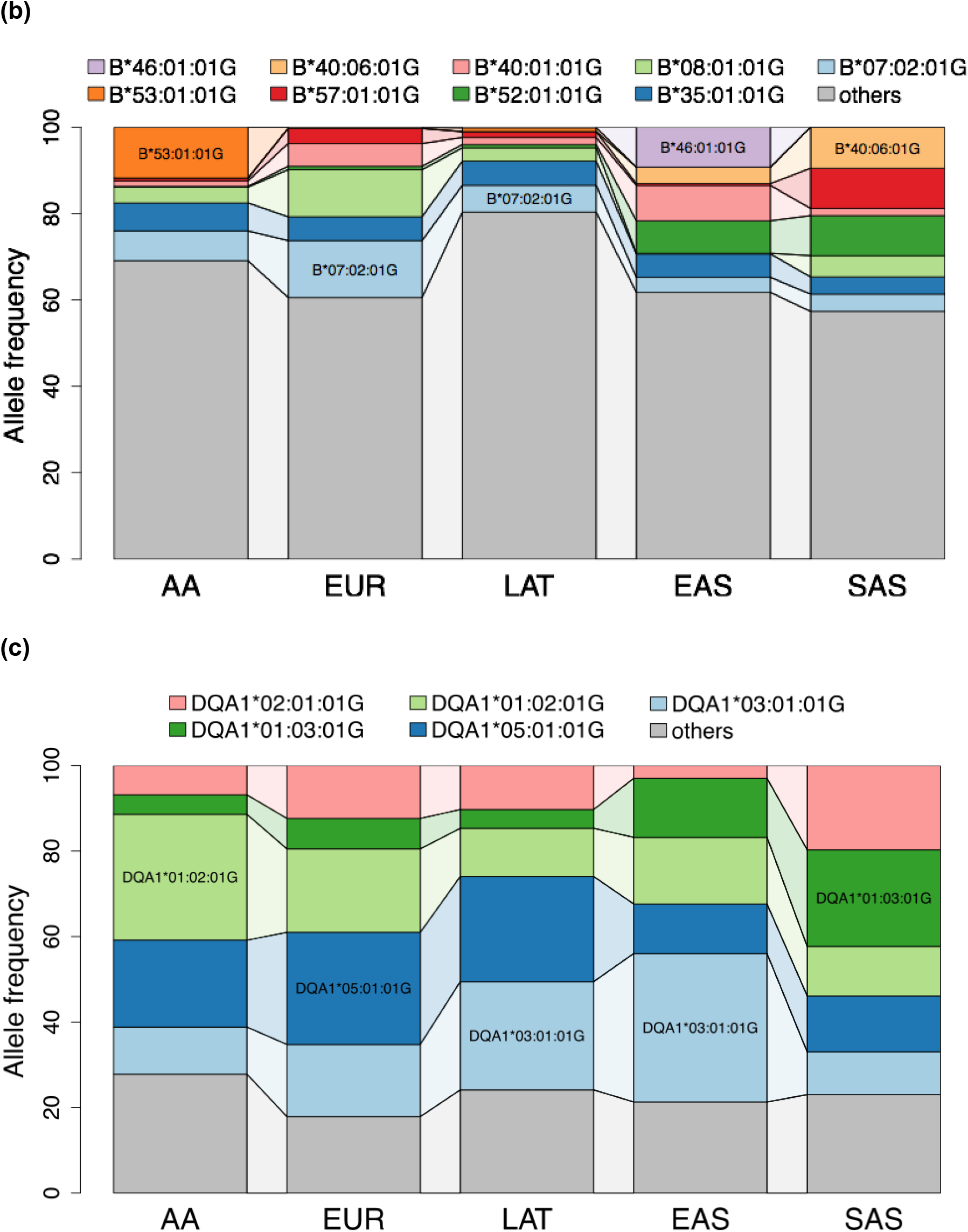
Global diversity of the MHC region. (**a**) Principal component analysis of the pairwise IBD distance between 21,546 samples using MHC region markers. Allele diversity of (**b**) HLA*-B* and (**c**) HLA-*DQA1* among five continental populations (AA=Admixed African; EUR=European; LAT=Latino; EAS=East Asian; SAS=South Asian). The top two most common alleles within each population group are named, the remaining alleles are grouped as ‘others’.

### HLA diversity

To quantify MHC diversity, we calculated identity-by-descent (IBD) distances^26^ between all individuals using 38,398 MHC single nucleotide polymorphisms (SNPs) included in the multi-ethnic HLA reference panel (N=21,546) and applied principal component analysis (PCA, **Methods**). PCA distinguished EUR, EAS and AA as well as the admixed LAT and SAS samples (**Figure 1a, Supplementary Figure 2**). This reflected widespread *HLA* allele frequency differences between populations (**Figure 1b-c, Supplementary Figure 3**). Of 130 unique common (frequency > 1%) G-group alleles, 129 demonstrated significant differences of frequencies across populations (4 degree-of-freedom Chi-square test, p-value < 0.05/130, **Supplementary Figure 4**). The only exception was *DQA1*01:01:01G* which was nominally significant (unadjusted p-value = 0.047). These differences may be related to adaptive selection. For example, the *B*53:01:01G* allele is enriched in Admixed Africans (11.7% in AA versus 0.3% in others) and it has been previously associated with malaria protection^27,28^. Consistent with previous reports^29,30^, we observed that HLA*-B* had the highest allelic diversity (n=443) while HLA-*DQA1* had the least (n=17, **Supplementary Figure 5-6, Supplementary Table 6, Extended Data 1**).

**Figure 2.**
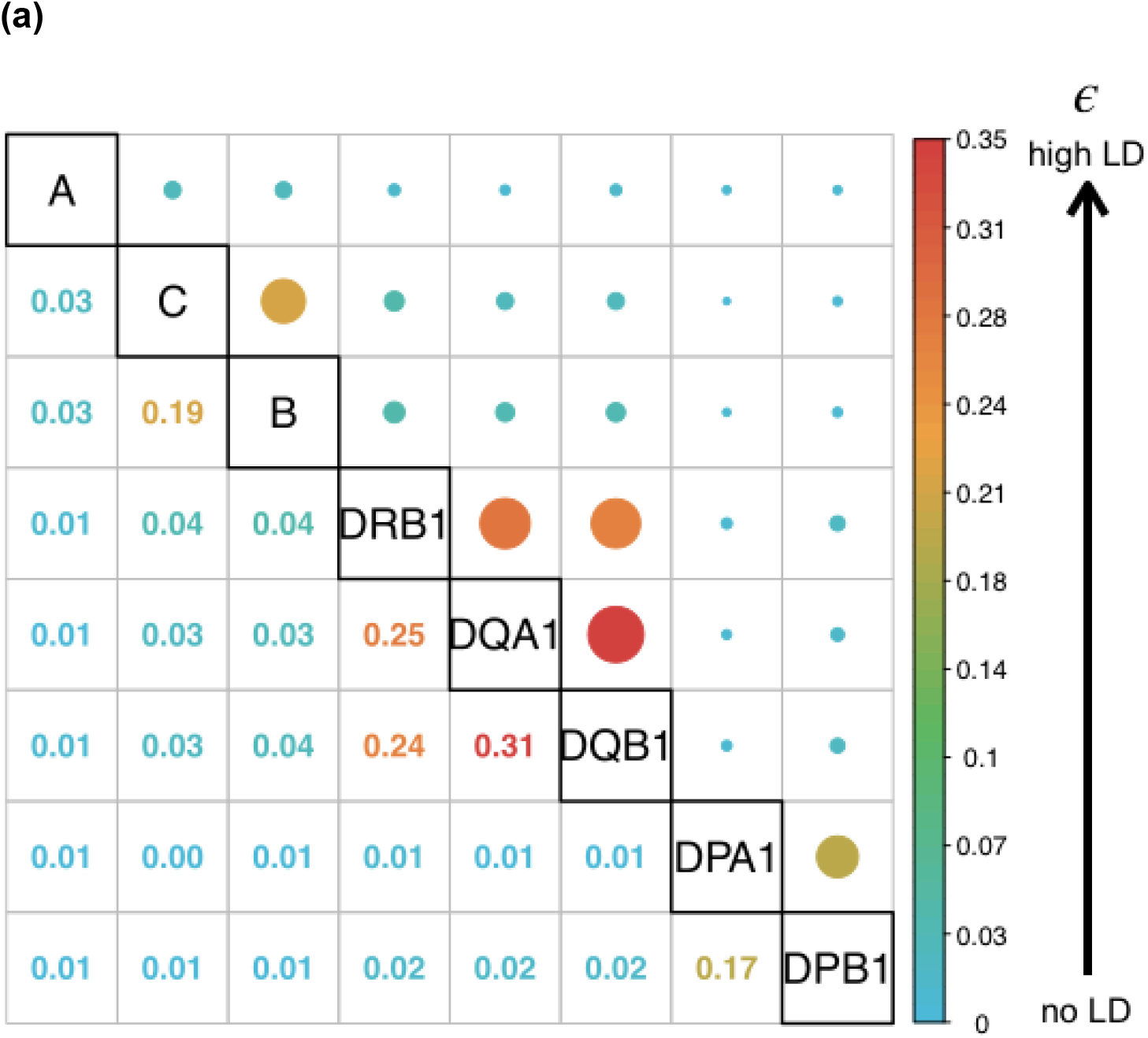
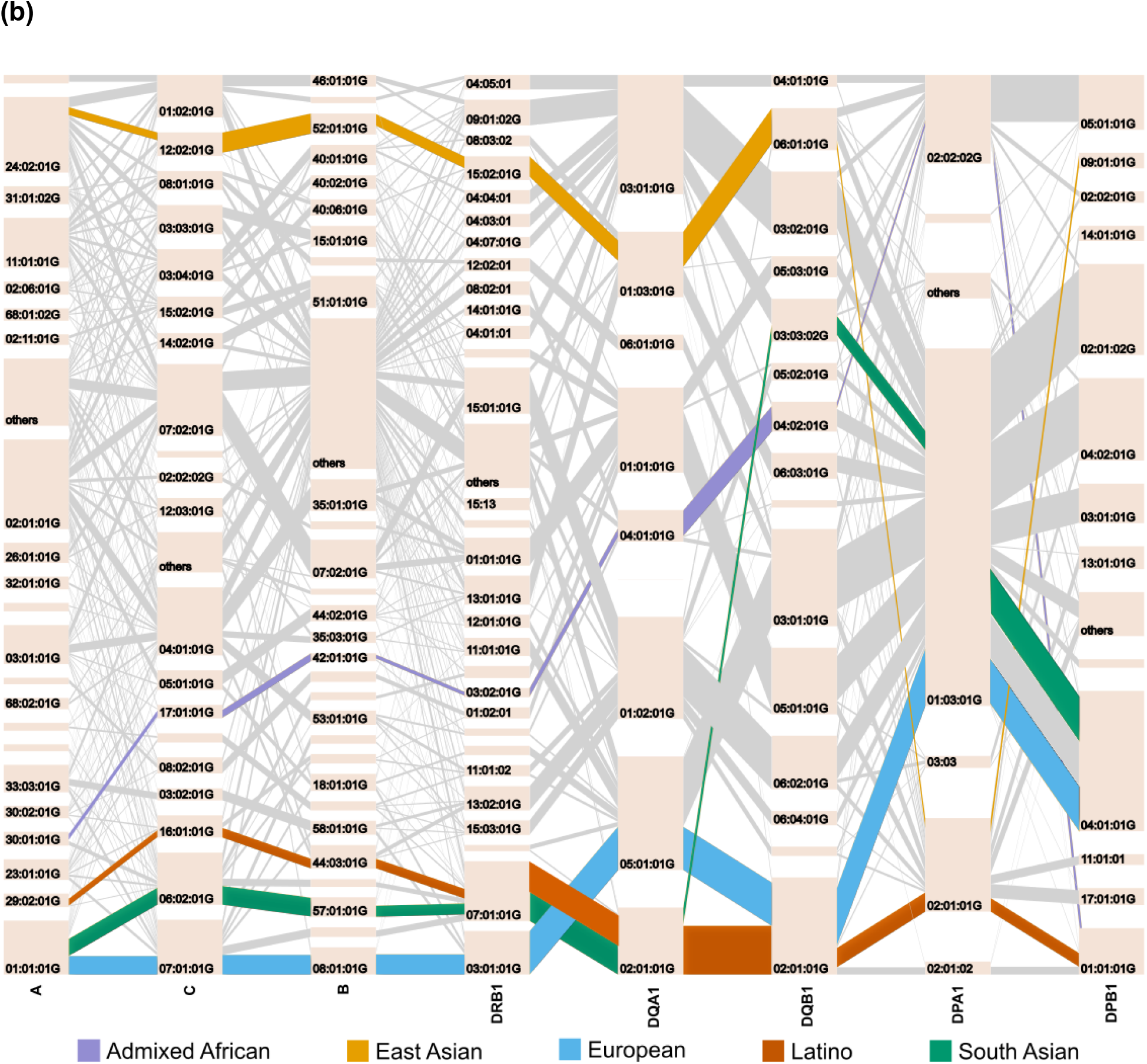
Pairwise LD and haplotype structure for six classical HLA genes in five population groups. (**a**) shows the pairwise normalized entropy (ε) measuring the difference of the haplotype frequency distribution for linkage disequilibrium and linkage equilibrium among five population groups. It takes values between 0 (no LD) to 1 (perfect LD). (**b**) shows the haplotype structures of the eight classical HLA genes in each population. The tile in a bar represents an *HLA* allele, and its height corresponds to the frequencies of the *HLA* allele. The gray lines connecting between two alleles represent *HLA* haplotypes. The width of these lines corresponds to the frequencies of the haplotypes. The most frequent long-range HLA haplotypes within each population is bolded and highlighted in a color described by the key at the bottom.

**Figure 3.**
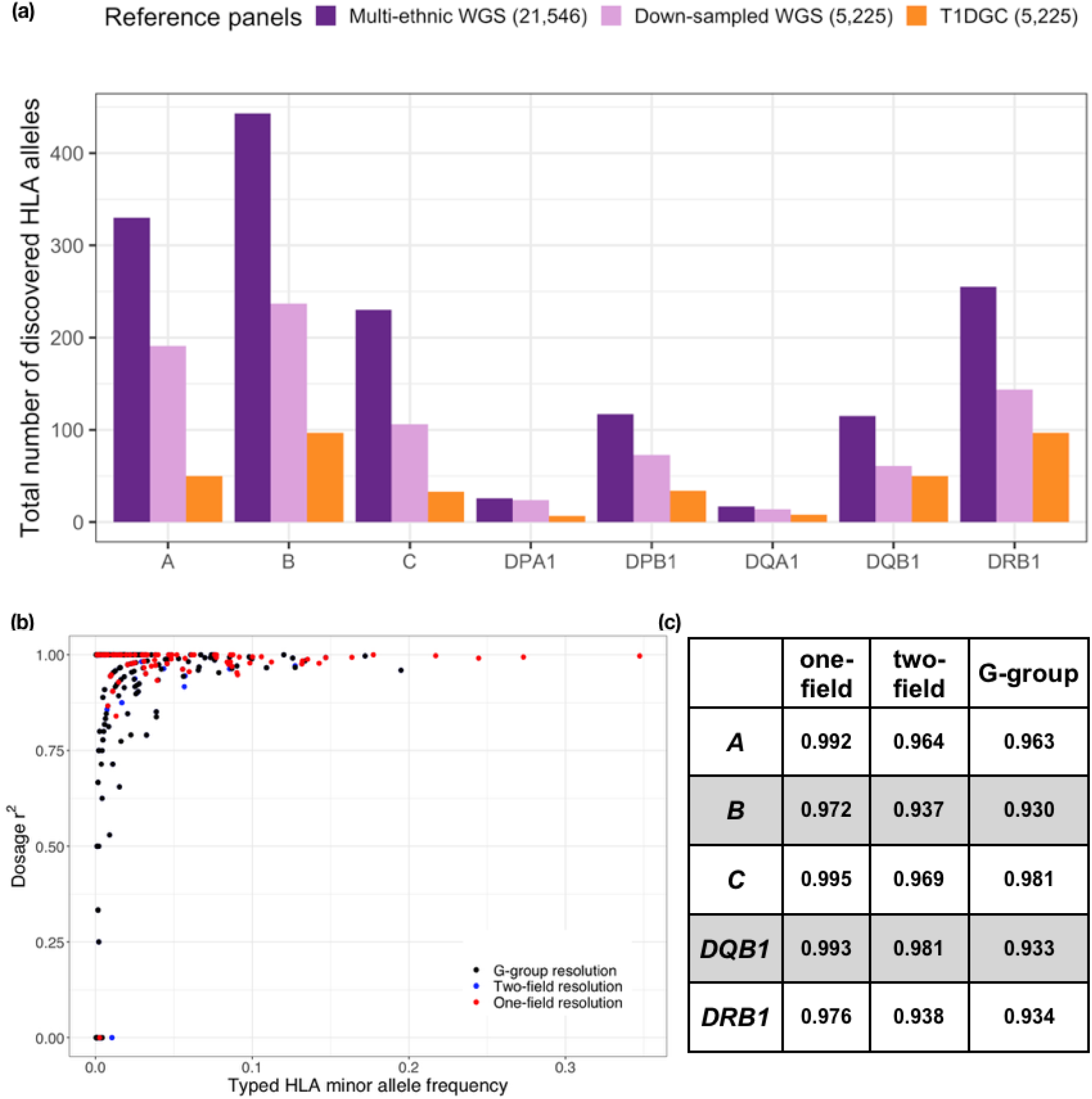
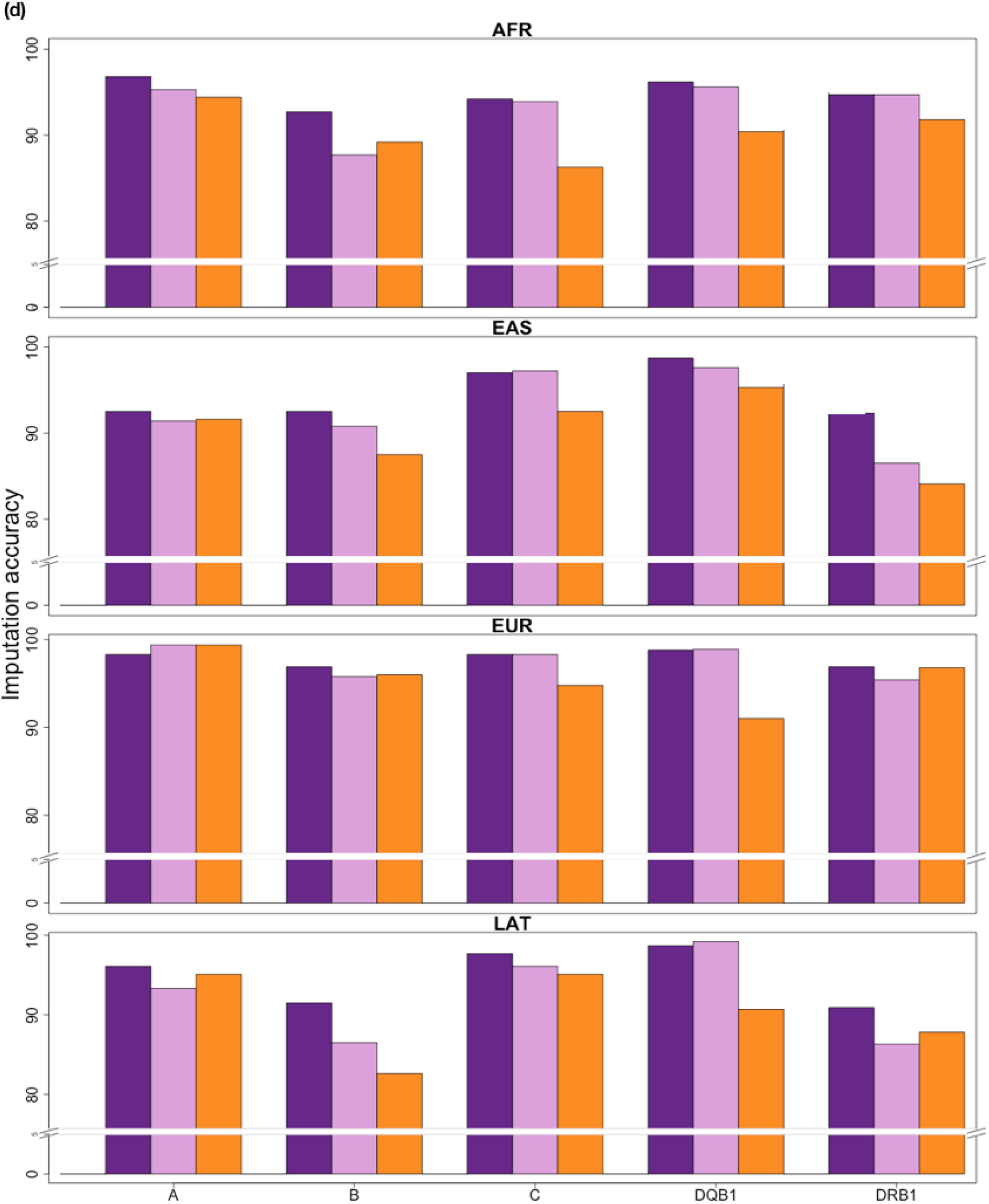
The multi-ethnic HLA reference panel shows improvement in allele diversity and imputation accuracy. (**a**). The number of HLA alleles at the two-field resolution included in the multi-ethnic HLA reference panel (N = 21,546) compared to the European only Type 1 Diabetes Genetics Consortium^48^ (T1DGC) panel (N = 5,225) as well as a subset of the multi-ethnic HLA panel down-sampled to the same size as T1DGC. (**b)**. The correlation between imputed and typed dosages of classical *HLA* alleles using the multi-ethnic HLA reference panel at one-filed (red), two-field (blue) and G-group resolution (black) of the 955 1000 Genomes subjects. (**c**). The imputation accuracy for five classical HLA genes at one-field, two-field and G-group resolution. (**d**). The imputation accuracy at G-group resolution of the 1000 Genomes subjects stratified by four diverse ancestries when using three different imputation reference panels as described in (**a**).

**Figure 4.**
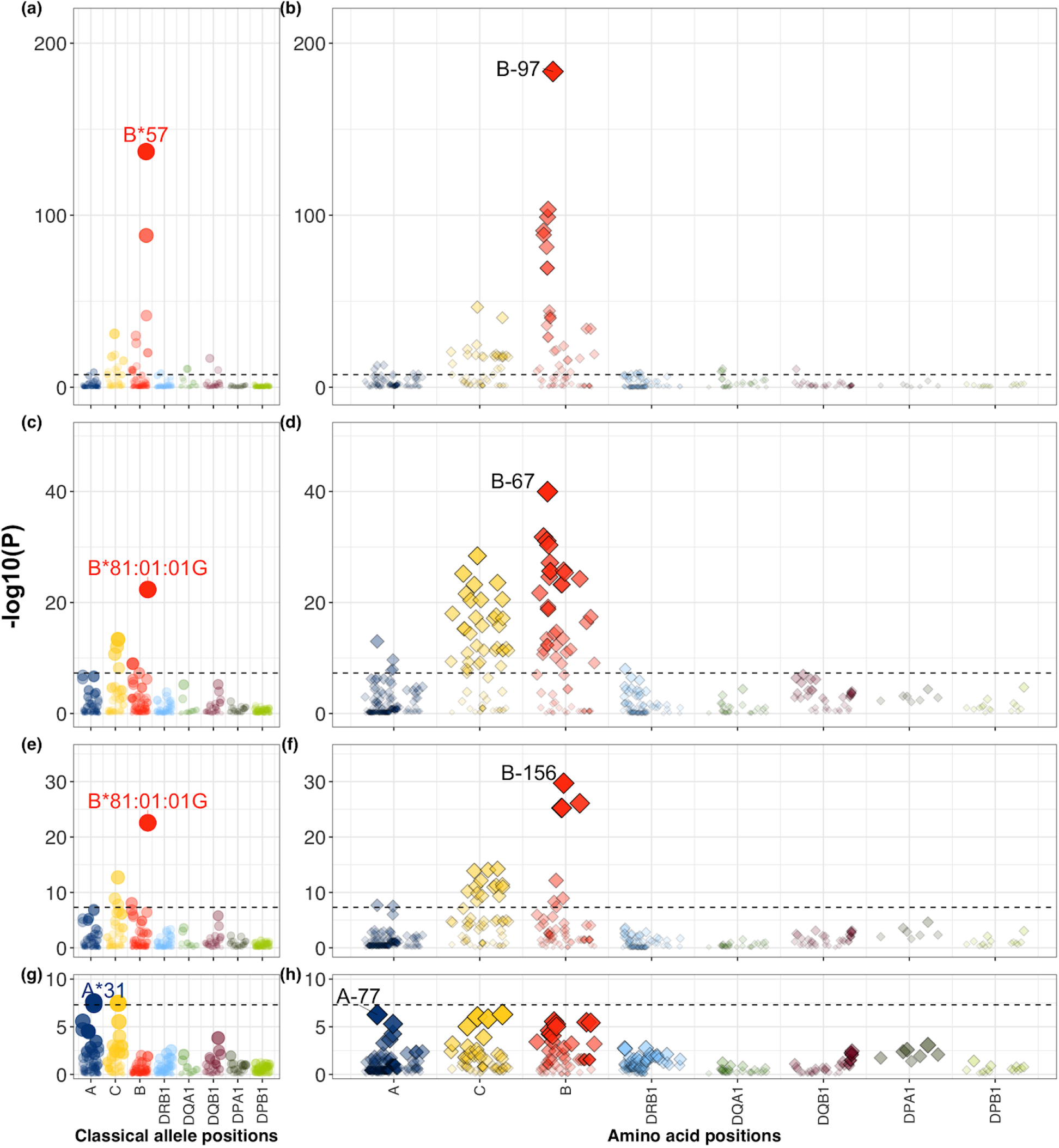
Stepwise conditional analysis of the allele and amino acid positions of classical HLA genes to HIV-1 viral load. Each circle point represents the linear regression -log10(*P*_*binary*_) for all classical *HLA* alleles. Each diamond point represents -log10(*P*_*omnibus*_) for the tested amino acid positions in HLA (blue=HLA-*A*; yellow=HLA-*C*; red=HLA-*B*; lightblue=HLA-*DRB1*; green=HLA*-DQA1*; purple=HLA-*DQB1*, darkgreen=HLA-*DPA1*; lightgreen=HLA-*DPB1*). Association at amino acid positions with more than two alleles was calculated using a multi-degree-of-freedom omnibus test. The dashed blacked line represents the significance threshold of *P* = 5 × 10^−8^. Each panel shows the association plot in the process of stepwise conditional omnibus test. (**a**) One-field classical allele *B*57* (*P* = 9.84 × 10^−138^) and (**b**) amino acid position 97 in HLA-B (*P* _*omnibus*_= 2.86 × 10^−184^) showed the strongest association signal. Results conditioned on position 97 in HLA-B showed a secondary signal at (**c**) classical allele *B*81:0101:G* (*P* = 4.53 × 10^−23^) and (**d**) position 67 in HLA-B (*P* _*omnibus*_= 1.08 × 10^−40^). Results conditioned on position 97 and 67 in HLA-B showed the same classical allele (**e**) *B*81:0101G* (*P* = 2.70 × 10^−23^) and (**f**) third signal at position 156 in HLA-B (*P* _*omnibus*_= 1.92 × 10^−30^). Results conditioned on position 97, 67 and 156 int HLA-B showed a fourth signal at (**g**) HLA*-A*31* (*P* = 2.45 × 10^−8^) and (**h**) position 77 in HLA-A (*P*_*omnibus*_ = 5.35 × 10^−7^) outside HLA-B.

**Figure 5.**
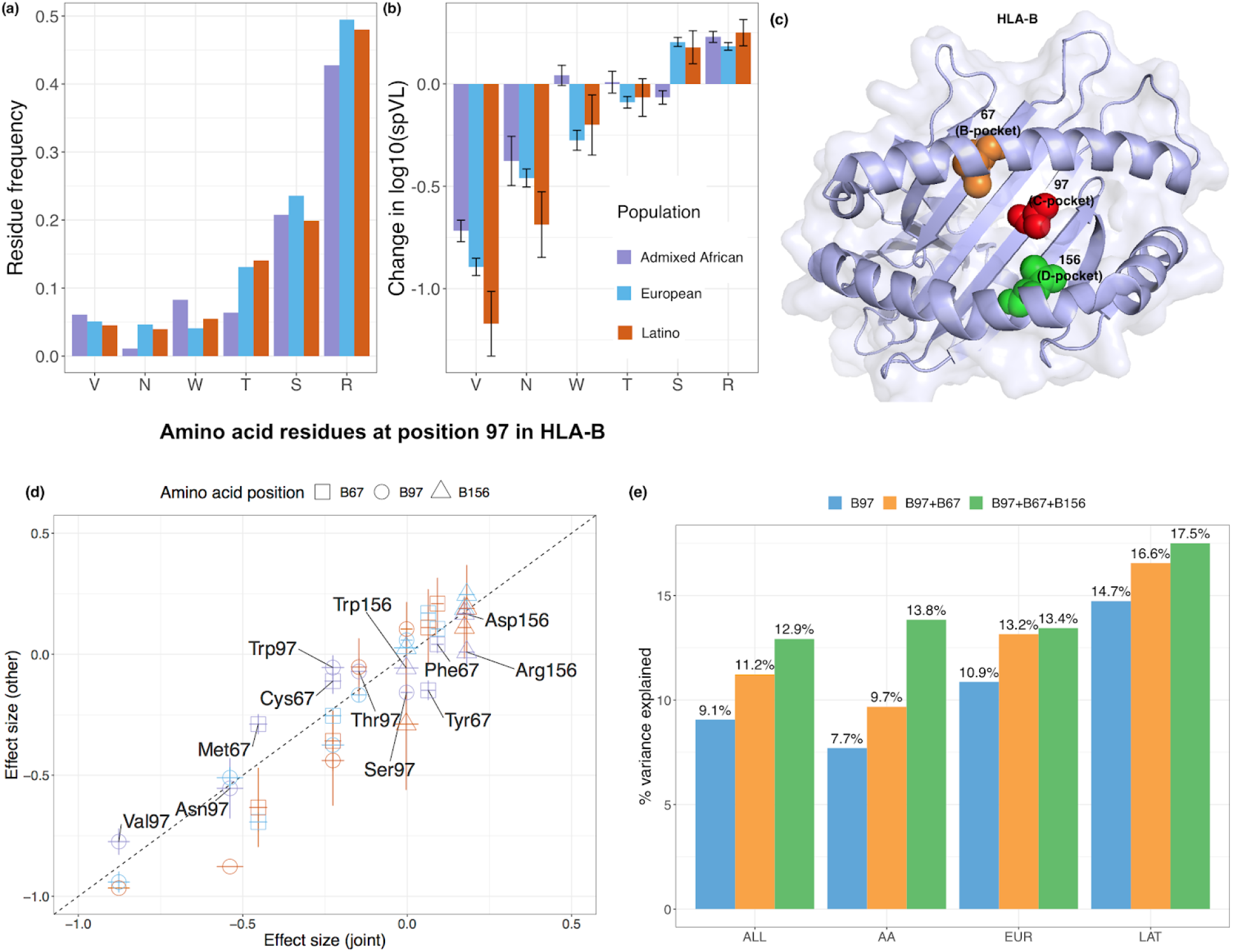
Location and effect of three independently associated amino acid positions in HLA-B. (**a**) Allele frequency of six residues at position 97 in HLA-B among three populations. (**b**) Effect on spVL (i.e., change in log10 HIV-1 spVL per allele copy) of individual amino acid residues at position 97 in HLA-B. Results were calculated per allele using linear regression models, including gender and principal components within each ancestry as covariates. (**c**) HLA-B (PDB ID code 2bvp) proteins. Omnibus and stepwise conditional analysis identified three independent amino acid positions (positions 97 (red), 67 (orange), and 156 (green) in HLA-B. (**d**) Effect on spVL (i.e., change in log10 HIV-1 spVL per allele copy) of individual amino acid residues at each position reported in this and previous work^10,16^. Results were calculated per allele using linear regression models. The x-axis shows the effect size and its standard errors in the joint analysis, and the y-axis shows the effect size and its standard error in individual populations (purple = Admixed American; blue = European and orange = Latino). (**e**) Variance of spVL explained by the haplotypes formed by different amino acid positions.

To understand the haplotype structure of HLA between pairs of HLA genes we calculated a multiallelic linkage disequilibrium (LD) measurement index^31–33^, ε, which is 0 when there is no LD and 1 when there is perfect LD (**Figure 2a**). We observed higher ε between *DQA1, DQB1*, and *DRB1;* between *DPA1* and *DPB1*; and between *B* and *C* (**Supplementary Figure 7**). The heterogeneity between different populations was underscored by the presence of population-specific common (frequency >1%) high resolution long-range haplotypes (HLA-*A∼C∼B∼DRB1∼DQA1∼DQB1∼DPA1∼DPB1*, **Figure 2b, Supplementary Figure 8-12, Extended Data 2, Methods**). The most common within-population haplotype was A24::DP6 (HLA*-A*24:02:01G∼C*12:02:01G∼B*52:01:01G∼DRB1*15:02:01G∼DQA1*01:03:01G∼DQB1*06 :01:01G∼DPA1*02:01:01G∼DPB1*09:01:01G*) found at a frequency of 3.61% in EAS (**Supplementary Figure 8**). This haplotype is strongly associated with immune-mediated traits such as HIV^34^ and ulcerative colitis^35^ in Japanese individuals. The next most common haplotype was the well-described European-specific ancestral haplotype A1::DP1 or 8.1^36,37^ (frequency=2.76%,

HLA-*A*01:01:01G∼C*07:01:01G∼B*08:01:01G∼DRB1*03:01:01G∼DQA1*05:01:01G∼DQB1*02: 01:01G∼DPA1*02:01:02G∼DPB1*01:01:01G*, **Supplementary Figure 9**). This haplotype is associated with diverse immunopathological phenotypes in the European population, including systemic lupus erythematosus^38^, myositis^39^ and several other conditions^36^. We observed long-range haplotypes in admixed populations including A1::DP4 in SAS (frequency=1.86%,

**Supplementary Figure 10**), A30::DP1 in AA (frequency=1.18%,HLA*-A*30:01:01G∼C*17:01:01G∼B*42:01:01:G∼DRB1*03:02:01G∼DQA1*04:01:01G∼DQB1*04 :02:01G∼DPA1*02:02:02G∼DPB1*01:01:01G*, **Supplementary Figure 11**), and A29::DP11 in LAT (frequency=0.74%,

HLA*-A*29:02:01G∼C*16:01:01G∼B*44*03:01:G∼DRB1*07:01:01G∼DQA1*02:01:01G∼DQB1*0 2:01:01G∼DPA1*02:01:01G∼DPB1*11:01:01G*, **Supplementary Figure 12**).

These haplotypes also have associations with multiple diseases: for example *C*06:02∼B*57:01* is associated with psoriasis^40^ and *A*30:01∼C*17:01∼B*42:01* is associated with HIV^41^.

### HLA selection signature

Previous studies have suggested that recent natural selection favors African ancestry in the HLA region in admixed populations^42–45^. To test this hypothesis in our data, we obtained WGS data from a subset of individuals within two admixed populations (1,832 AA and 594 LAT, determined by the first three global principal components, **Supplementary Figure 13, Supplementary Note**). Admixed individuals have genomes that are a mosaic of different ancestries. If genetic variations or haplotypes from an ancestral population are advantageous, then they are under selection and are expected to have higher frequency than by chance. Using ELAI^46^, we quantified how much the ancestry proportions differed within the MHC from the genome-wide average. In AA, we observed that the average genome-wide proportion of African ancestry was 74.5%, compared to 78.0% in the extended MHC region, corresponding to a 3.42 (95% CI: 3.35-3.49) standard deviation increase. In LAT, we observed 5.76% African ancestry genome-wide versus 16.0% in the extended MHC region, representing an increase of 4.23 (95% CI: 4.14-4.31) standard deviations (**Methods, Supplementary Figure 14**). To ensure our results are robust to different local ancestry inference methods, we applied an alternative method called RFMix^47^ and observed a similarly consistent MHC-specific excess of African ancestry in LAT, and also an excess in AA that was more modest (**Supplementary Figure 14)**.

### Construction of a multi-ethnic HLA reference panel and its performance evaluation

Next, we constructed a multi-ethnic HLA imputation reference panel based on classical HLA alleles and 38,398 genomic markers in the extended MHC region using a novel HLA-focused pipeline HLA-TAPAS (HLA-Typing At Protein for Association Studies). Briefly, HLA-TAPAS can handle HLA reference panel construction (*MakeReference*); HLA imputation (*SNP2HLA*) and HLA association (*HLAassoc*) (**Methods, URLs**). Compared to a widely used HLA reference panel with European-only individuals (The Type 1 Diabetes Genetics Consortium^48^, T1DGC), this new reference panel has a six-fold increase in the number of observed *HLA* alleles and non-HLA genomic markers (**Supplementary Table 7**). We noted the difference in observed classical *HLA* alleles is mainly due to the inclusion of diverse populations rather than its size; after downsampling the reference panel to be the same size as T1DGC (N=5,225), there was still a three-fold increase in observed alleles (**Figure 3a**).

To empirically assess imputation accuracy of our reference panel, we first used the publicly available gold-standard *HLA* types (HLA-*A*, -*B*,-*C*, -*DRB1* and -*DQB1*) of 1,267 diverse samples from AA, EAS, EUR and LAT included in 1KG. We removed 955 overlapping samples within the reference panel, and to ensure a representative analysis we kept 6,007 markers overlapping with the *Global Genotyping Array* SNPs. Across the five genes, the average G-group resolution accuracies were 94.2%, 93.7%, 97.8% and 93.7% in AA, EAS, EUR and LAT (**Figure 3b-c, Supplementary Table 8, Methods, Extended Data 3**). Compared to the T1DGC panel, our multi-ethnic reference panel showed the most improvement for individuals of non-European descent; we obtained 4.27%, 2.96%, 2.90% and 1.05% improvement at G-group resolution for AA, EAS, LAT, and EUR individuals, respectively (**Figure 3d**). Increased diversity was responsible for the improvement; downsampling the reference panel be the same size as the T1DGC panel still yielded superior performance (**Figure 3d**). To validate our panel further, we imputed *HLA* alleles into a multi-ethnic cohort of 2,291 individuals from the Genotype and Phenotype (GaP) registry genotyped on the ImmunoChip array. We obtained SBT *HLA* type information for six classical class I and class II loci (HLA-*A*, -*B*, -*C*, -*DQA1*, -*DQB1*, -*DRB1*) in 75 samples with diverse ancestral background (25 EUR, 25 EAS and 25 AA, **Supplementary Figure 15, Methods**). Average accuracies were 99.0%, 95.7% and 97.0% for EUR, EAS and AA respectively when comparing SBT *HLA* alleles at G-group resolution (**Methods, Extended Data 3**). Similar to the 1KG analysis, the multi-ethnic reference panel showed significant improvement for individuals with non-European descent (6.3% and 11.1% improvement for EAS and African individuals respectively at G-group resolution), and a more modest 2% improvement in EUR (**Supplementary Figure 16, Supplementary Table 9**).

### Fine-mapping causal variants of HIV jointly in three populations in the MHC region

Next we investigated MHC effects within human immunodeficiency virus type 1 (HIV-1) set point viral load. Upon primary infection with HIV-1, the set point viral load is reached after the immune system has developed specific cytotoxic T lymphocytes (CTL) that are able to partially control the virus. It has been well-established that the set point viral load (spVL) varies in the infected population and positively correlates with rate of disease progression^49^. Previous studies suggested that HIV-1 infection has a strong genetic component, and specific HLA class I alleles explain the majority of genetic risk^10,50^. The existence of multiple independent, ancestry-specific, risk-associated alleles has been reported in both European^1,10^ and African American^16^ populations. However, without a multi-ethnic reference panel it has not been possible to determine if these signals are consistent across different ancestral groups.

To define the MHC allelic effects shared across multiple populations, we applied our multi-ethnic MHC reference panel to 7,445 EUR, 3,901 AA and 677 LAT HIV-1 infected subjects (**Methods, Supplementary Table 10**). Imputation resulted in 640 classical HLA alleles, 4,513 amino acids in HLA proteins and 49,321 SNPs in the extended MHC region for association and fine-mapping analysis. We confirmed 96.6% imputation accuracy of two-field (or four-digit) resolution with a minor allele frequency > 0.5% in this cohort by comparing imputed classical alleles to the SBT alleles in a subset of 1,067 AA subjects^16^(**Supplementary Figure 17, Extended Data 3**).

We next tested SNPs, amino acid positions and classical *HLA* alleles across the MHC for association to spVL. We performed this jointly in EUR, AA and LAT population using a linear regression model with sex, principal components and ancestry as covariates (**Methods**). In agreement with previous studies, we found the strongest spVL-associated classical *HLA* allele is *B*57* (effect size = −0.84, *P*_*binary*_ = 8.68 × 10^−144^). This corresponded to a single residue Val97 in HLA-B that tracks almost perfectly with *B*57* (*r*^2^ = 0.995) and showed the strongest association of any single residue (effect size = −0.84, *P*_*binary*_ = 5.99 × 10^−145^, **Supplementary Figure 18**).

Then to determine which amino acid positions have independent association with spVL, we tested each of the amino acid positions by grouping haplotypes carrying a specific residue at each position in an additive model^2,9^ (**Methods**). We found the strongest spVL-associated amino acid variant in HLA-B is as previously reported^1,10,16^ at position 97 (**Figure 4a-b, Supplementary Table 11**) which strikingly explains 9.06% of the phenotypic variance. Position 97 in HLA-B was more significant (*P*_*omnibus*_ = 2.86 × 10^−184^) than any single SNP or classical *HLA* allele, including *B*57* (**Supplementary Figure 18, Extended Data 4**). Of the six allelic variants (Val/Asn/Trp/Thr/Arg/Ser) at this position, the Val residue conferred the strongest protective effect (effect size = −0.88, *P* = 9.32 × 10^−152^, **Supplementary Figure 19**) relative to the mos common residue Arg (frequency = 47.8%). All six amino acid alleles have consistent frequencies and effect sizes across the three population groups (**Figure 5a-b, Supplementary Figure 20**).

We next wanted to test whether there were other independent effects outside of position 97 in HLA-B. After accounting for the effects of amino acid 97 in HLA-B using a conditional haplotype analysis (**Methods**), we observed a significant independent association at position 67 in HLA-B (*P*_*omnibus*_ = 2.82 × 10^−39^, **Figure 4c-d, Supplementary Table 11**). Considering this might be an artifact of forward search, we exhaustively tested all possible pairs of polymorphic amino acid positions in HLA-B. Of 7,260 pairs of amino acid positions, none obtained a better goodness-of-fit than the pair of positions 97 and 67, which collectively explained 11.2% variance in spVL (**Figure 5e, Supplementary Table 12**). At position 67, Met67 residue shows the most protective effect (effect size = −0.44, *P* = 1.19 × 10^−59^) among the five possible amino acids (Cys/Phe/Met/Ser/Tyr) relative to the most common residue Ser (frequency =10.0%).

Conditioning on positions 97 and 67 revealed an additional association at position 156 in HLA-B (*P*_*omnibus*_ = 1.92 × 10^−30^, **Figure 4e-f, Supplementary Table 11**). In agreement with the stepwise conditional analysis, when we tested all 287,980 possible combinations of three amino acid positions in HLA-B, the most statistically significant combination of amino acids sites is 67, 97 and 156 (*P* = 5.68 × 10^−244^, **Supplementary Table 13**). These three positions explained 12.9% of the variance (**Figure 5e**). At position 156, residue Arg shows the largest risk effect (effect size = 0.180, *P* = 8.92 × 10^−14^) among the four possible allelic variants (Leu/Arg/Asp/Trp), relative to the most common residue Leu (frequency = 35.1%).

These amino acid positions mark three consecutive pockets within the HLA-B peptide-binding groove (**Figure 5c**). Position 97 is located in the C-pocket and has an important role in determining the specificity of the peptide-binding groove^51,52^. Position 67 is in the B-pocket, and Met67 side chains occupy the space where larger B-pocket anchors reside in other peptide-MHC structures; its presence limits the size of potential peptide position P2 side chains^52^. Amino acid position 156 is part of the D-pocket and influences the conformation of the peptide-binding region^53^. These results are consistent with the observation that in HLA-*B*57*, the single most protective spVL-associated one-field allele (a single change at position 156 from Leu → Arg or equivalently HLA-*B*57:03 →* HLA-*B*57:02*) leads to an increased repertoire of HIV-specific epitope^41,54^.

Despite differences in the power to detect associations due to differences in allele frequencies (**Supplementary Figure 21**), we observed generally consistent effects of individual residues across populations (**Figure 5d, Supplementary Figure 22-23, Supplementary Table 14**). There are 26 unique haplotypes defined by the amino acids at positions 67, 97 and 156 in HLA-B (**Table 1, Supplementary Table 15**). When we tested for effect size heterogeneity by ancestry for each of these haplotypes (**Methods**), we observed only 2 of 26 haplotypes showed heterogeneity (F-test P-value < 0.05/26), possibly due to different interplay between genetic and environmental variation at population-level. These results support the concept that these positions mediate HIV-1 viral load in diverse ancestries.

To assess whether there were other independent MHC associations outside HLA-B, we conditioned on all amino acid positions in HLA-B and observed associations at HLA-A, including at position 77 in HLA-A (*P* _*omnibus*_= 9.10 × 10^−7^, **Figure 4g-h, Supplementary Table 11**), the classical *HLA* allele *HLA-A*31* (*P* _*binary*_= 2.45 × 10^−8^) and the *rs2256919* promoter SNP (*P*_*binary*_ =3.10 × 10^−16^, **Supplementary Figure 18**). These associations argue for an effect at HLA-A, but larger studies and functional studies will be necessary to define the driving effects.

## Discussion

In our study we demonstrated accurate imputation with a single large reference panel for HLA imputation. We have shown how this reference panel can be used to impute genetic variation at eight *HLA* classical genes accurately across a wide range of populations. Accurate imputation in multi-ethnic studies is essential for fine-mapping.

We showed the utility of this approach by defining the alleles that best explain HIV-1 viral load in infected individuals. Our work implicates three amino acid positions (97, 67 and 156) in HLA-B in conferring the known protective effect of HLA class I variation on HIV-1 infection. Combining all alleles at these three positions explained 12.9% of the variance in spVL (**Figure 5e**). These positions all fall within the peptide-binding groove of the respective MHC protein (**Figure 5c**), indicating that variation in the amino acid content of the peptide-binding groove is the major genetic determinant of HIV control. Supported by experimental studies^54–57^, positions highlighted in our work indicated a structural basis for the HLA association with HIV disease progression that is mediated by the conformation of the peptide within the class I binding groove. This result highlights how a study with ancestrally diverse populations can potentially point to causal variation by leveraging linkage disequilibrium difference between ethnic groups.

We note that previous studies have shown position 97 in HLA-B has the strongest association with HIV-1 spVL or case-control in African American and European populations, but highlighted different additional signals via conditional analysis (position 45, 67 in HLA-B and position 77, 95 in HLA-A in Europeans^1,10,16^ and position 63, 116 and 245 in HLA-B in African Americans^16^). These signals do not explain the signals we report here; after conditioning on positions 45, 63, 116, 245 of HLA-B and 95 of HLA-A, the association of the four identified amino acids identified in this study remained significant (*P* < 5 × 10^−8^). In contrast, our binding groove alleles explain these other alleles; conditioning on the four amino acid positions identified in this study (positions 67, 97 and 156 in HLA-B), all previously reported positions did not pass the significance threshold (*P* > 5 × 10^−8^, **Supplementary Figure 24**).

Furthermore, defining the effect sizes for *HLA* alleles across different populations is essential for defining risk of a wide-range of diseases in the clinical setting. There is increasing application of genome-wide genotyping by patients both by healthcare providers and direct-to-consumer vendors. The large effects of the MHC region for a wide-range of immune and non-immune traits, makes it essential to define *HLA* allelic effect sizes essential in multi-ethnic studies in order to build generally applicable clinical polygenic risk scores for many diseases in diverse populations^58–61^. Resources like the one we present here will be an essential ingredient in such studies.

## Methods

### Individuals included in the reference panel

Study participants were from the Jackson Heart Study (JHS, N = 3,027), Multi-Ethnic Study of Atherosclerosis (MESA, N=4,620), Chronic Obstructive Pulmonary Disease Gene (COPDGene) study (N=10,623), Estonian Biobank (EST, N=2,244), Japan Biological Informatics Consortium (JPN, N=295), Biobank Japan (JPN, N=1,025) and 1000 Genomes Project (1KG, N=2,504). Each study was previously approved by respective institutional review boards (IRBs), including for the generation of WGS data and association with phenotypes. All participants provided written consent. Further details of cohort descriptions and phenotype definitions are described in the **Supplementary Note**.

### HLA-TAPAS

HLA-TAPAS (HLA-Typing At Protein for Association Studies) is an HLA-focused pipeline that can handle HLA reference panel construction (*MakeReference*), HLA imputation (*SNP2HLA*), and HLA association (*HLAassoc*). It is an updated version of the SNP2HLA^48^ to build an imputation reference panel, perform *HLA* classical allele, amino acid and SNP imputation within the extended MHC region. Briefly, major updates include (1) using PLINK1.9 (**URLs**) instead of v1.07; (2) using BEAGLE v4.1 (**URLs**) instead of v3 for phasing and imputation; and (3) including custom R scripts for performing association and fine-mapping analysis at amino acid level in multiple ancestries. The source code is available for download (**URLs**).

### Construction of a multi-ethnic HLA reference panel using whole-genome sequences

To construct a multi-ethnic HLA imputation reference panel, we used 24,338 whole-genome sequences at different depths (**Supplementary Table 1**). Details of the construction using deep-coverage whole-genome sequencing are described in the **Supplementary Note**. Briefly, alignment and variant-calling for genomes sequenced by each cohort were performed independently. We performed local realignment and quality recalibration with the Genome Analysis Toolkit^62^ (GATK; version 3.6) on Chromosome 6:25,000,000-35,000,000. We detected single nucleotide variants (SNV) and indels using GATK with HaplotypeCaller. To eliminate false-positive sites called in the MHC region, we restrict our panel to SNVs reported in 1000 Genomes Project^21^ only.

We next inferred classical HLA alleles at G-group resolution for eight classical HLA genes (*HLA-A*, -*B*, -*C*, -*DQA1*, -*DQB1*, -*DRB1, -DPA1* and -*DPB1*) using a population reference graph^24,25^. To extend the reference panel versatility, we inferred amino acid variation, one-field and two-field resolution alleles from the inferred G-group alleles. After removing samples with low-coverage and failed genome-wide quality control (**Supplementary Table 3**), we constructed a multi-ethnic HLA imputation reference panel (N=21,546) using the HLA-TAPAS *MakeReference* module (**URLs, Method**).

### Sequence-based typing of *HLA* alleles

Purified DNA from the 75 donors from the GaP registry (at the Feinstein Institute for Medical Research) was sent to NHS Blood and Transplant, UK, where *HLA* typing was performed. Next-generation sequencing was done for HLA*-A, -B, -C*, -*DQB1*, -*DPB1* and *-DRB1*.

PCR-sequence-specific oligonucleotide probe sequencing was performed for HLA*-DQA1* in all samples. These typing methods yielded classical allele calls for seven genes at three-field (HLA-*A*, -*B*, -*C* and -*DQB1*) or G-group resolution (HLA-*DQA1*, -*DPB1* and *-DRB1*).

Genomic DNA from the 288 unrelated samples of Japanese ancestry underwent high-resolution allele typing (three-field alleles) of six classical HLA genes (HLA-*A*, -*B* and -*C* for class I; and HLA-*DRB1*, -*DQA1* and -*DPB1* for class II)^20^.

The 1000 Genomes panel consists of 1,267 individuals with information on five HLA genes (HLA-*A, -B, -C, -DQB1*, and *-DRB1*) at G-group resolution among four major ancestral groups (AA, EAS, EUR and LAT)^7^.

We obtained HLA typing of the 1,067 African American subjects included in the HIV-1 viral load study as described previously^16,63^. Briefly, seven classical HLA genes (HLA-*A, -B, -C, -DQA1*, -*DQB1* -*DRB1* and *-DPB1*) were obtained by sequencing exons 2 and 3 and/or single-stranded conformation polymorphism PCR, and was provided at two-field resolution.

### Accuracy measure between inferred and sequence-based typing *HLA* genotypes

Allelic variants at HLA genes can be typed at different resolutions: one-field HLA types specify serological activity, two-field HLA types specify the amino acids encoded by the exons of the HLA gene, and three-field types determine the full exonic sequence including synonymous variants. G-group resolution determines the sequences of the exons encoding the peptide binding groove, that is, exons 2 and 3 for class I and exon 2 class II genes. Thus, any polymorphism occurring in exon 4 of class I gene or exon 3 of class II gene was not defined. This means many G-group alleles can map to multiple three-field and two-field *HLA* alleles.

We calculated the accuracy at each *HLA* gene by summing across the dosage of each correctly inferred *HLA* allele or amino acid across all individuals (N), and divided by the total number of observations (2*N). That is,

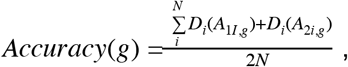

where *Accuracy*(*g*) represents the accuracy at a classical HLA gene (e.g. HLA-*B*). *D*_*i*_represents the inferred dosage of an allele in individual *i*, and alleles *A*_1*i,g*_ and *A*_2*i,g*_ represent the true (SBT) *HLA* types for an individual *i*.

To evaluate the accuracy between the inferred and validated *HLA* types obtained from SBT at G-group resolution, we translated the highest resolution specified by the validation data to its matching G-group resolution based IMGT/HLA database (e.g. HLA-*A*01:01* → HLA-*A*01:01:01G*), and compared it to the primary output from *HLA*LA* or *HLA-TAPAS*. We also translated all G-group alleles to their matching amino acid sequences, and compared them against the validation alleles, we referred to this as the amino acid level.

To evaluate imputation performance in individual classical *HLA* alleles and amino acids, we calculated the dosage *r*^2^ correlation between imputed and SBT dosage.

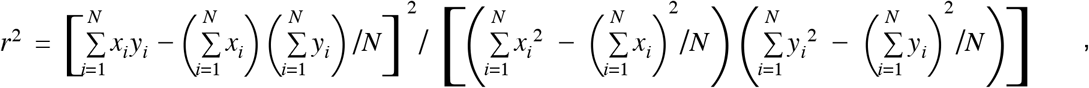

where *x*_*i*_ and *y*_*i*_ represents the inferred and SBT dosage of an allele in individual *i*. *N* represents the number of individuals.

### Principal component analysis

We performed a principal component analysis of the MHC region based on the identity-by-descent (IBD) distances between all 21,809 individuals included in the multi-ethnic reference panel. We computed the IBD distance using Beagle (Version 4.1, **URLs**) and averaged over 100 runs with all variants (54,474) included in the HLA reference panel. Due to uneven representation of different ethnicity groups (**Supplementary Table 2**), we applied a weighted PCA approach, where mean and standard deviation of the IBD matrix within an ethnicity group are weighted inversely proportional to the sample size.

### HLA haplotype frequency estimation

We applied an expectation-maximization algorithm approach implemented in Hapl-o-Mat^64^ (**URLs**) to estimate HLA haplotype frequency based on eight classical HLA alleles inferred at G-group resolution. We estimated haplotype frequencies both overall and within five continental populations (**Extended Data 2**).

### Local ancestry inference

To detect local ancestry in admixed samples, we first applied ELAI^46^ to chromosome 6 with 1000 Genomes Project^21^ as the reference panel. We extracted 63,998 common HapMap3 SNPs between the WGS (MESA cohort) and the 1000 Genome reference panel. We used the same set of SNPs for ELAI and RFMix analysis. We applied ELAI^46^ to 1,832 African Americans and 594 Latinos. For 1,832 African American individuals included in the study, we used genotypes of 99 CEU and 108 YRI in the 1000 Genome Project as reference panel, assuming admixture generation to be seven generations ago. We used two upper-layer clusters and 10 lower-layer clusters in the model. For Latinos, we selected 65 Latinos with Native American (NAT) ancestry > 75% included in the 1000 Genomes Project identified using the ADMIXTURE analysis^65^ and used these individuals with high NAT, as well as CEU and YRI from 1000 Genomes as reference panels. We assumed that the admixture time was 20 generations ago. For ELAI, we used three upper-layer clusters and 15 lower-layer clusters in the model.

To address the technical concerns that local ancestry methods are biased by the high LD of MHC region^66,67^, we performed an alternative method, RFMix^47^, for local ancestry inference that accounts for high LD and lack of parental reference panels. Similar deviation from genome-wide ancestry was observed using RFMix (**Supplementary Figure 14)**, indicating that the selection signals we observed here are robust to different inference methods.

### HLA imputation in the HIV-1 viral load GWAS data in three populations

We used genome-wide genotyping data from 12,023 HIV-1 infected individuals aggregated across more than 10 different cohorts (**Supplementary Table 10**). The details of these samples and quality control procedures have been described previously^10,68^. Using the HIV-1 viral load GWAS data, we extracted the genotypes of SNPs located in the extended MHC region (chr6:28-34Mb, **Supplementary Table 10**). We conducted genotype imputation of one-field, two-field and G-group classical *HLA* alleles and amino acid polymorphisms of the eight class I and class II HLA genes using the constructed multi-ethnic HLA imputation reference panel and the HLA-TAPAS pipeline.

After imputation, we obtained the genotypes of 640 classical alleles, 4,513 amino acid positions of the eight classical HLA genes, and 49,321 SNPs located in the extended MHC region. We excluded variants with MAF < 0.5% and imputation *r*^2^ < 0.5 for all association studies. In total, we tested 51,358 variants in our association and fine-mapping study.

### HLA association analysis

For the HIV-1 viral loads of EUR, AA and LAT samples, we conducted a joint haplotype-based association analysis using a linear regression model under the assumption of additive effects of the number of HLA haplotypes for each individual. Phased haplotypes at a locus (i.e., HLA amino acid position) were constructed from the phased imputed genotypes of variants in the locus (i.e., amino acid change or SNP) and were converted to a haplotype matrix where each row is observed haplotypes (in the locus), not genotypes.

For each amino acid position, we applied a conditional haplotype analysis. We tested a multiallelic association between the HIV-1 viral load and a haplotype matrix (of the position) with covariates, including sex, study-specific PCs, and a categorical variable indicating a population. That is

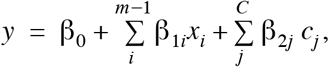

where *x*_*i*_ is the amino acid haplotype formed by each of the *m* amino acid residues that occur at that position, and *c*_*j*_ are the covariates included in the model.

To get an omnibus *P*-value for each position, we estimated the effect of each amino acid by assessing the significance of the improvement in fit by calculating the in-model fit, compared to a null model following an F-distribution with degrees of freedom. This is implemented using an ANOVA test in R as described previously^32,69^. The most frequent haplotype was excluded from a haplotype matrix as a reference haplotype for association.

**For the conditional analysis**, we assumed that the null model consisted of haplotypes as defined by residues at previously defined amino acid positions. The alternative model is in addition of another position with *m* residues. We tested whether the addition of those amino acid positions, and the creation of additional haplotypes groups, improved on the previousset. We then assessed the significance of the improvement in the delta deviance (sum of squares) over the previous model using an F-test. We performed stepwise conditional analysis to identify additional independent signals by adjusting for the most significant amino acid position in each step until none met the significance threshold (*P* = 5 × 10^−8^). We restricted analysis to haplotypes that have a minimum of 10 occurrences within HLA-B, and removed any individual with rare haplotypes for the conditional analysis.

**For the exhaustive search**, we tested all possible amino acid pairs and triplets for association. For each set of amino acid positions, we used the groups of residues occurring at these positions to estimate effect size and calculated for each of these models the delta deviance in risk prediction and its p-values compared to the null model.

### Heterogeneity testing of effect sizes

We used interaction analyses with models that included haplotype-by-ancestry (*Haplotype × Ancestry*) interaction terms. The fit of nested models was compared to a null model using the *F*-statistic with two degrees of freedom, for which the association interaction P-value indicated whether the inclusion of the *Haplotype × Ancestry* interaction terms improved the model fit compared to the null model that did not include the interaction terms. Interaction P-values for all haplotypes formed by positions 97, 67 and 156 in HLA-B are listed in **Supplementary Table 15**. Haplotypes that had a significant Bonferroni-corrected *Haplotype × Ancestry* interaction heterogeneity P-value (P < 0.05/26) were considered to show evidence of significant effect size heterogeneity between ancestries.

## Data Availability

The source code is available for download at https://github.com/immunogenomics

## URLs

HLA-TAPAS, https://github.com/immunogenomics/HLA-TAPAS

IMGT/HLA, https://www.ebi.ac.uk/ipd/imgt/hla/;

GATK version 3.6, https://software.broadinstitute.org/gatk/download/archive;

HLA*LA, https://github.com/DiltheyLab/HLA-PRG-LA;

PLINK 1.90, https://www.cog-genomics.org/plink2;

Beagle 4.1, https://faculty.washington.edu/browning/beagle/b4_1.html;

Hapl-o-Mat, https://github.com/DKMS/Hapl-o-Mat/;

1000 Genomes gold-standard HLA types, http://ftp.1000genomes.ebi.ac.uk/vol1/ftp/data_collections/HLA_types/

## Acknowledgements

The study was supported by the National Institutes of Health (NIH) TB Research Unit Network, Grant U19 AI111224-01.

The views expressed in this manuscript are those of the authors and do not necessarily represent the views of the National Heart, Lung, and Blood Institute; the National Institutes of Health; or the U.S. Department of Health and Human Services.

The Genotype and Phenotype (GaP) Registry at The Feinstein Institute for Medical Research provided fresh, de-identified human plasma; blood was collected from control subjects under an IRB-approved protocol (IRB# 09-081) and processed to isolate plasma. The GaP is a sub-protocol of the Tissue Donation Program (TDP) at Northwell Health and a national resource for genotype-phenotype studies.

https://www.feinsteininstitute.org/robert-s-boas-center-for-genomics-and-human-genetics/gap-registry/ A.M. is supported by Gentransmed grant 2014-2020.4.01.15-0012.; D.W.H. is supported by NIH grants AI110527, AI077505, TR000445, AI069439, and AI110527. D.H.S. was supported by R01 HL92301, R01 HL67348, R01 NS058700, R01 AR48797, R01 DK071891, R01 AG058921, the General Clinical Research Center of the Wake Forest University School of Medicine (M01 RR07122, F32 HL085989), the American Diabetes Association, and a pilot grant from the Claude Pepper Older Americans Independence Center of Wake Forest University Health Sciences (P60 AG10484). J.T.E. and P.E.S. were supported by NIH/NIAMS R01 AR042742, R01 AR050511, and R01 AR063611.

For some HIV cohort participants, DNA and data collection was supported by NIH/NIAID AIDS Clinical Trial Group (ACTG) grants UM1 AI068634, UM1 AI068636 and UM1 AI106701, and ACTG clinical research site grants A1069412, A1069423, A1069424, A1069503, AI025859, AI025868, AI027658, AI027661, AI027666, AI027675, AI032782, AI034853, AI038858, AI045008, AI046370, AI046376, AI050409, AI050410, AI050410, AI058740, AI060354, AI068636, AI069412, AI069415, AI069418, AI069419, AI069423, AI069424, AI069428, AI069432, AI069432, AI069434, AI069439, AI069447, AI069450, AI069452, AI069465, AI069467, AI069470, AI069471, AI069472, AI069474, AI069477, AI069481, AI069484, AI069494, AI069495, AI069496, AI069501, AI069501, AI069502, AI069503, AI069511, AI069513, AI069532, AI069534, AI069556, AI072626, AI073961, RR000046, RR000425, RR023561, RR024156, RR024160, RR024996, RR025008, RR025747, RR025777, RR025780, TR000004, TR000058, TR000124, TR000170, TR000439, TR000445, TR000457, TR001079, TR001082, TR001111, and TR024160.

Molecular data for the Trans-Omics in Precision Medicine (TOPMed) program was supported by the National Heart, Lung and Blood Institute (NHLBI). See the TOPMed Omics Support Table (**Supplementary Table 16**) for study specific omics support information. Core support including centralized genomic read mapping and genotype calling, along with variant quality metrics and filtering were provided by the TOPMed Informatics Research Center (3R01HL-117626-02S1; contract HHSN268201800002I). Core support including phenotype harmonization, data management, sample-identity QC, and general program coordination were provided by the TOPMed Data Coordinating Center (R01HL-120393; U01HL-120393; contract HHSN268201800001I). We gratefully acknowledge the studies and participants who provided biological samples and data for TOPMed.

The COPDGene project was supported by Award Number U01 HL089897 and Award Number U01 HL089856 from the National Heart, Lung, and Blood Institute. The content is solely the responsibility of the authors and does not necessarily represent the official views of the National Heart, Lung, and Blood Institute or the National Institutes of Health. The COPDGene project is also supported by the COPD Foundation through contributions made to an Industry Advisory Board comprised of AstraZeneca, Boehringer Ingelheim, GlaxoSmithKline, Novartis, Pfizer, Siemens and Sunovion. A full listing of COPDGene investigators can be found at: http://www.copdgene.org/directory

The Jackson Heart Study (JHS) is supported and conducted in collaboration with Jackson State University (HHSN268201800013I), Tougaloo College (HHSN268201800014I), theMississippi State Department of Health (HHSN268201800015I) and the University of Mississippi Medical Center (HHSN268201800010I, HHSN268201800011I and HHSN268201800012I) contracts from the National Heart, Lung, and Blood Institute (NHLBI) and the National Institute on Minority Health and Health Disparities (NIMHD). The authors also wish to thank the staffs and participants of the JHS.

MESA and the MESA SHARe project are conducted and supported by the National Heart, Lung, and Blood Institute (NHLBI) in collaboration with MESA investigators. Support for MESA is provided by contracts 75N92020D00001, HHSN268201500003I, N01-HC-95159, 75N92020D00005, N01-HC-95160, 75N92020D00002, N01-HC-95161, 75N92020D00003, N01-HC-95162, 75N92020D00006, N01-HC-95163, 75N92020D00004, N01-HC-95164, 75N92020D00007, N01-HC-95165, N01-HC-95166, N01-HC-95167, N01-HC-95168, N01-HC-95169, UL1-TR-000040, UL1-TR-001079, UL1-TR-001420. MESA Family is conducted and supported by the National Heart, Lung, and Blood Institute (NHLBI) in collaboration with MESA investigators. Support is provided by grants and contracts R01HL071051, R01HL071205, R01HL071250, R01HL071251, R01HL071258, R01HL071259, by the National Center for Research Resources, Grant UL1RR033176. The provision of genotyping data was supported in part by the National Center for Advancing Translational Sciences, CTSI grant UL1TR001881, and the National Institute of Diabetes and Digestive and Kidney Disease Diabetes Research Center (DRC) grant DK063491 to the Southern California Diabetes Endocrinology Research Center. This project has been funded in whole or in part with federal funds from the Frederick National Laboratory for Cancer Research, under Contract No. HHSN261200800001E. The content of this publication does not necessarily reflect the views or policies of the Department of Health and Human Services, nor does mention of trade names, commercial products, or organizations imply endorsement by the U.S. Government. This Research was supported in part by the Intramural Research Program of the NIH, Frederick National Lab, Center for Cancer Research.

## Author contributions

Y. L. and S.R. conceived, designed and performed analyses, wrote the manuscript and supervised the research. M.K. implemented the omnibus test for the HIV-1 fine-mapping study. Y.L., W.C., M.K., P.E.S., J.T.E., and B.H. contributed to the development of the HLA-TAPAS pipeline. X.L. performed the selection analysis. J.T.E, M.G.-A. and P.K.G helped with the GaP data acquisition. K.Y., K.O., D.W.H., X.G., N.D.P., Y.I.C., J.I.R., K.D.T., S.S.R., A.C., J.G.W., S.K., M.H.C., A.M., T.E., and Y.O. contributed to the WGS data acquisition. J.F., M.C. and P.J.M contributed to the HIV-1 data acquisition. All authors contributed to the writing of the manuscript.

